# Human adenovirus outbreak at a university campus monitored by wastewater and clinical surveillance

**DOI:** 10.1101/2024.03.27.24304990

**Authors:** Steven C. Holland, Matthew F. Smith, LaRinda A. Holland, Rabia Maqsood, James C. Hu, Vel Murugan, Erin M. Driver, Rolf U. Halden, Efrem S. Lim

## Abstract

Areas of dense population congregation are prone to experience respiratory virus outbreaks. We monitored wastewater and clinic patients for the presence of respiratory viruses on a large, public university campus. Campus sewer systems were monitored in 16 locations for the presence of viruses using next generation sequencing over 22 weeks in 2023. During this period, we detected a surge in human adenovirus (HAdV) levels in wastewater. Hence, we initiated clinical surveillance at an on-campus clinic from patients presenting with acute respiratory infection. From whole genome sequencing of 123 throat and/or nasal swabs collected, we identified an outbreak of HAdV, specifically of HAdV-E4 and HAdV-B7 genotypes overlapping in time. The temporal dynamics and proportions of HAdV genotypes found in wastewater were corroborated in clinical infections. We tracked specific single nucleotide polymorphisms (SNPs) found in clinical virus sequences and showed that they arose in wastewater signals concordant with the time of clinical presentation, linking community transmission of HAdV to the outbreak. This study demonstrates how wastewater-based epidemiology can be integrated with surveillance at ambulatory healthcare settings to monitor areas prone to respiratory virus outbreaks and provide public health guidance.

**Graphical Abstract:** 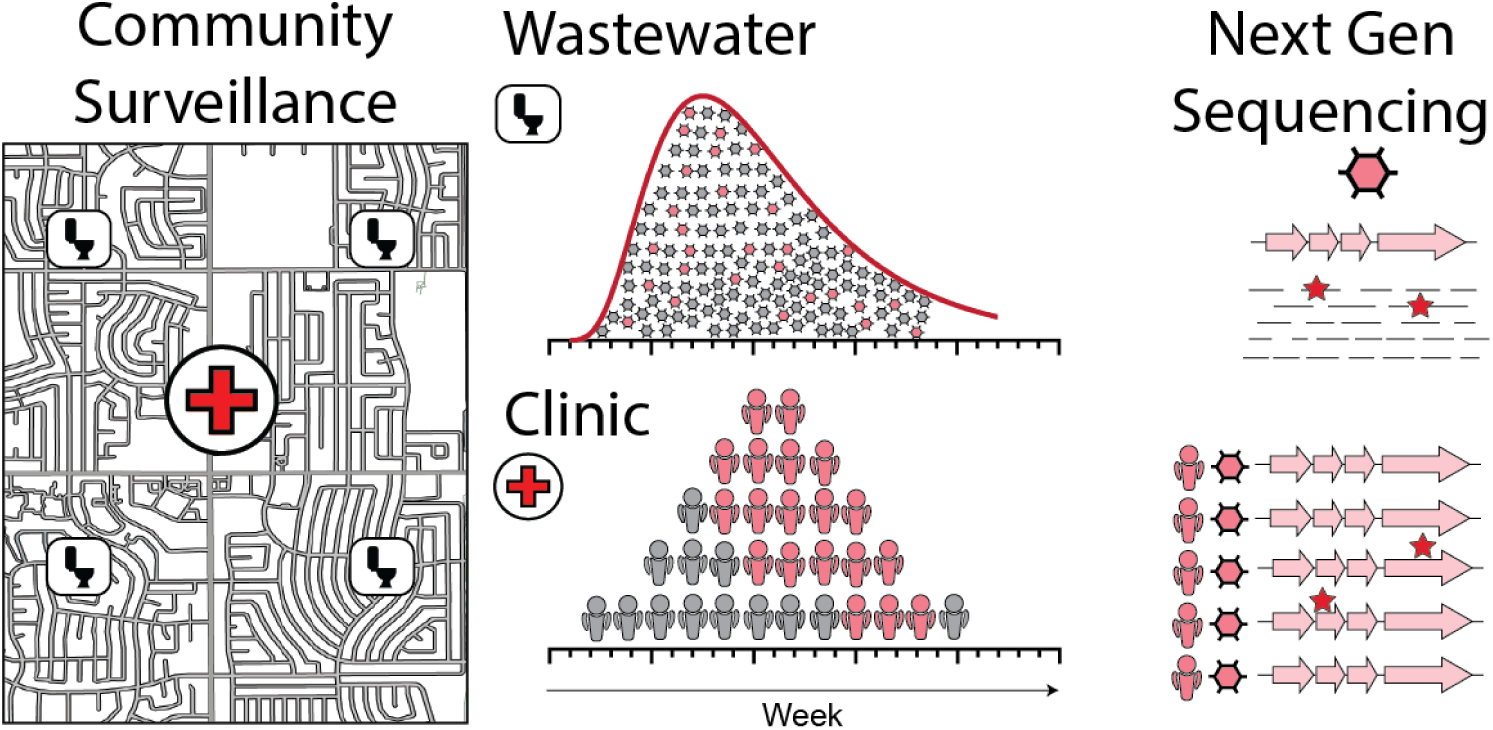

## 1. Introduction

Wastewater surveillance of biological pathogens, or wastewater-based epidemiology (WBE), has become an increasingly important framework for monitoring pathogen presence, evolution, and providing public health guidance (Prado et al., 2021; Sims and Kasprzyk-Hordern, 2020). WBE allows for interrogation of community health in an economical, scalable, and anonymous way. Traditionally WBE has been performed using PCR-based methods on single genetic biomarkers (Choi et al., 2018). Due to the improvement of molecular techniques to enrich genomes of interest and reduce assay inhibition, next generation sequencing across whole genomes from species present in wastewater at low-abundance is possible (Child et al., 2023). This enables improved public health monitoring of disease outbreaks and monitoring for immunologically relevant mutations of interest across whole genomes (Smith et al., 2023).

Appreciation for WBE as a public health surveillance tool is increasing as our understanding of how wastewater surveillance is also applicable to non-enteric pathogens, such as respiratory viruses (Boehm et al., 2023; Lowry et al., 2023). Respiratory viruses pose significant health risks, especially in infant and older adult populations (Boon et al., 2024; Goka et al., 2015). These viruses also present an economic burden due to health care costs and productivity losses (Fendrick et al., 2003). Since a concomitant rise in viral abundance in wastewater is observed during infectious disease outbreaks (Sims and Kasprzyk-Hordern, 2020), WBE can be used to detect outbreaks and inform public health guidance. Use of WBE in conjuction with clinical patient surveillance may further be able to inform therapeutic use, discriminate between genomic variants causing higher morbidity, and more clearly define transmission dynamics.

Human adenoviruses (HAdVs) are nonenveloped, linear double stranded DNA viruses endemic to most populations and occur without any strict seasonality (Moriyama et al., 2020). There are 7 major HAdV subtypes, A-G, each associated with different clinical manifestations and host age preferences. These clinical manifestations can be broad, ranging from respiratory distress (HAdV-B, -C, -E), gastroenteritis (HAdV-F), conjunctivitis (HAdV-D), myocarditis, haemorrhagic cystisis (HAdV-B) (Arnold and MacMahon, 2017). HAdV-F, an enteric HAdV, is ubiquitous in municipal wastewater, and may be used as an indicator of sewage contamination in the environment (Hewitt et al., 2013). Respiratory HAdVs (B, C, E) are still detectable in wastewater, but only a few studies investigated on how their abundance relates to disease prevalence (Lun et al., 2019). Although HAdV infections are often common and self-limiting, outbreaks can cause acute respiratory distress, particularly in congregated communities like universities and military training grounds (Cheng et al., 2016; Yusof et al., 2012). HAdVs are resistant to disinfectants and able to be spread by droplet inhalation, fomites, faecal-oral transmission, and the transplantation of tissues (Arnold and MacMahon, 2017).

In the present study we monitored a large, public U.S. university campus for the presence of respiratory viruses in wastewater sewer systems and from patients visiting an on-campus health clinic. We show that a surge of HAdV in wastewater coincided with an outbreak of HAdV infections presenting at the campus clinic as outpatients. We tracked unique genomic markers identified in clinical cases in the wastewater sequencing and observed that wastewater signals corroborated clinical presentation.

## 2. Materials and Methods

### 2.1. Wastewater collection and sample processing

Wastewater samples were collected weekly between January and May 2023 at manhole locations across the campus. Week of sample collection are according to the MMWR week numbering convention. Sites were selected with knowledge of the sewer infrastructure and chosen to isolate a single building or a collective of buildings. Moore’s swab passive samplers comprised of 2-3 cotton tampons housed inside an organza bag were deployed for a minimum of 24-hours at each collection site. Upon retrieval, samplers were stored in polyethene (PE) bags and transported on ice to the laboratory for same day processing. For each sampler, the edge of the PE bag was clipped and the contents of the sampler squeezed onto a 0.45 µM polyethersulfone vacuum filtration unit (Fisherbrand, Waltham, MA). The filtrate was transferred to a 15mL Amicon® Ultra Centrifugal Filter with 10 kDa molecular weight cut off (Millipore Sigma, Burlington, MA) for concentration with five rounds of centrifugation at 3700 rpm for 15-20 minutes at 4 °C. Subsequently, 200 µL of each sample concentrate were extracted using a Qiagen RNeasy Mini Kit (Hilden, Germany) to a final volume of 50 µL. Additional method details can be found in previous work (Bowes et al., 2023; Wright et al., 2022).

### 2.2. Wastewater viral quantification

SARS-CoV-2 viral load in wastewater samples was quantified using a 3-gene multiplexed qRT-PCR assay previously described (Fontenele et al., 2023). Briefly, the Applied BiosystemsTM TaqPathTM COVID-19 Combo Kit and TaqPathTM Multiplex Master Mix (No ROX) were used (Thermo Fisher Scientific - Waltham, MA), with the following modifications per reaction well: 10 μL nuclease free water, 6.25 μL multiplex master mix, 2.5 μL MS2 phage control (diluted 1:10 with nuclease free water), and 1.25 μL COVID-19 real-time PCR assay multiplex, 5 μL of sample. Thermal conditions were as follows: hold for 2 min at 25 °C, 10 min at 53 °C, 2 min at 95 °C, followed by 40 cycles of 95 °C for 3 s and 60 °C for 30 s.

HAdV viral load in wastewater samples was quantified using HAdV-B3 and HAdV-B7 specific primer and probes sequences obtained from a previously published assay. (Lu et al., 2013). In our hands, their HAdV-E4 primer/probe set was unsuccessful, so used the HAdV-E4 primer/probe sequences from another published assay. (Lion et al., 2003). Final HAdV-B3 qPCR reactions contained 300 nmol forward primer (P_f_), 600 nmol reverse primer (P_r_), and 200 nmol probe. Final HAdV-B7 qPCR reactions contained 600 nmol Pf, 300 nmol Pr, and 50 nmol probe. Final HAdV-E4 qPCR reactions contained 300 nmol Pf, 600 nmol Pr, and 200 nmol probe. All reactions contained 1X TaqMan Fast Universal PCR master mix, no AmpErase (Applied Biosystems, Waltham, MA), 5 µL of wastewater TNA extract, and water to a final volume of 25 µL. Reaction conditions were 95°C for 1 min, followed by 40 cycles of (i) 95°C for 15 s, (ii) 60°C for 30 s. A previously published peppered mild mottle virus assay was used as a control (Haramoto et al., 2013). Reactions contained 400 nmol Pf, 400 nmol Pr, and 200 nmol probe, 1 X SuperScript III One-Step Reaction mix (Thermo Fisher, Waltham, MA), 1 X SuperScript III RT/Platinum Taq mix (Invitrogen, Waltham, MA), 5 µL of waster TNA extract, and water to a final volume of 25 µL. Ten-fold serial dilutions using synthetic DNA fragments of primer/probe targets were used to generate were used to create standard curves to quantify genome copies and asses qPCR efficiency (**Supplementary Figs. S1A, S1B, & S1C**.

### 2.3. Clinical collection

Symptomatic individuals seeking care for an acute respiratory infection of ≤ 7 days duration at the University’s Health Services clinic were recruited in this study. Patients with illness duration >7 days at the time of respiratory specimen collection were excluded from the study. Study participants provided informed written or verbal consent for study enrollment. The study was approved by site reliance on Duke University and Arizona State University Institutional Review Boards. Respiratory specimens (nasopharyngeal swabs and throat swabs combined) were collected from clinical patients and stored at 4°C until nucleic acid extraction.

### 2.4. Nucleic acid extraction, hybrid enrichment, sequencing

Total nucleic acids of wastewater and clinical specimens were extracted using the KingFisher Flex System (Thermo Scientific, Waltham, MA) according to manufacturer guidelines. Libraries were prepared using the Illumina RNA Prep (Illumina, San Diego, CA) with enrichment using the Illumina Respiratory Virus Oligo Panel v2 (Illumina, San Diego, CA). Libraries were sequenced with Illumina 2 x 150 paired end reads (Illumina, San Diego, CA).

#### 2.4.1. Sequencing read mapping and consensus genome synthesis

Demultiplexed sequencing reads were trimmed of adapter sequences and removed of phiX and human genome contamination using bbtools v39 (Bushnell, 2014). Sequencing reads were mapped to genomes using the Burrows-Wheeler aligner v0.7.17-r1188(Li and Durbin, 2009). Consensus genomes were generated using samtools 1.17 (Li et al., 2009) at a minimum read depth ≥10 and quality ≥ 20. Samples with ≥ 500 reads mapping to a viral genome (after negative control subtraction) with genome coverage ≥ 25% were considered positive for an infection with that virus.

#### 2.4.2. Phylogenetics

Phylogenetic analysis was performed on clinical sample consensus sequences and sequences published in publicly available databases. For SARS-CoV-2, samples were analyzed using Nextstrain CLI version 8.2.0 (Hadfield et al., 2018) using the SARS-CoV-2 global open reference dataset and local surveillance references, subsampled to 38 total genomes with collection dates within the study period. HAdV references sequences were found by created a consensus genome sequence for each genotype and performed a BLAST search of the NIH Nucleotide collection (nr/nt) and compared that sequence to the top scoring result. The sequenced HAdV-B3 genome differed from a HAdV-B3 genome isolated in China during 2009 by 8 SNPs (MK836311). The consensus sequence of clinical B7 genomes differed by one synonymous SNP, a27797g, to an HAdV-B7 genome isolated from a child in China during 2019 (MT113942)(Wei et al., 2023). The consensus sequence of E4 genomes was identical to an HAdV-E4 genome found in the USA during a 2022 outbreak (OP785759)(Tori et al., 2024). Reference sequences for other genotypes were found by Genbank annotation. Full-length genomes from representative sub-clades were identified from recent phylogenetic analyses of coronaviruses (Shao et al., 2022; Woo et al., 2006), human metapnuemoviruses (Nao et al., 2020), human parainfluenza viruses (Linster et al., 2018; Shao et al., 2021), and obtained from Genbank. Representative genomes of influenza and human rhinovirus were selected by Genbank classification from major subfamilies. Consensus genomes from clinical specimens and Genbank genomes were aligned using MAFFT version v7.520 (Katoh et al., 2002) with default parameters. Maximum likelihood trees were created using IQTREE2 version v2.2.0.3 (Minh et al., 2020) with default arguments. Phylogenetics trees were visualized using FigTree version v1.4.4.

#### 2.4.3. Wastewater SNP analysis

Detection of HAdV polymorphisms in wastewater samples was performed by mapping wastewater reads to HAdV-B3, HAdV-B7, and HAdV-E4 consensus sequences from the clinical cohort. Wastewater sequencing reads were mapped to the genomes using the Burrows-Wheeler aligner v0.7.17-r1188. Variant call files were generated using samtools version v1.17 (Li et al., 2009) and ivar version v1.4.2 (Castellano et al., 2021). Only wastewater samples with variant frequency ≥ 0.005 and read depth ≥ 10 were reported.

#### 2.4.4. SARS-CoV-2 wastewater analysis

For wastewater samples achieving at least 50% breadth of SARS-CoV-2 genome coverage, lineage abundance was estimated using Freyja v1.4.9 (Karthikeyan et al., 2022) from wastewater sequencing reads successfully mapped to the Wuhan1 reference genome. For each sample, relative abundance of PANGO sublineages were aggregated to parental lineages. Specifically, BQ.1 and sublineages were aggregated to BQ.1, BA.2 and sublineages were aggregated to BA.2, XBB.1.5 and sublineages were aggregated to XBB.1.5, XBB.1.9 and sublineages were aggregated to XBB.1.9, XBB.2 and sublineages were aggregated to XBB.2, XBB.1 and sublineages were aggregated to XBB.1 except XBB.1.5 and XBB.1.9 sublineages, all other XBB sublineages except XBB.1.5, XBB.1.9, XBB.2, and XBB.1 sublineages were aggregated to Other XBB, all other recombinant X lineages were aggregated to Other Recombinants, and all other lineages were aggregated to Other. Aggregated abundances were averaged across samples within each week for visualization.

#### 2.4.5. Statistical analysis

Principle coordinate analysis was performed on wastewater relative abundance values using the phyloseq package v1.38.0 in R statistical software using the ordinate() command with method=”PCoA” and distance=”bray” arguments. Kendall’s Tau correlation tests with Benjamini-Hochberg multiple test correction was performed in R using the corr.test() function, method = “kendal”. Linear regression, and lognormal curve fittings were performed using GraphPad Prism (La Jolla, CA) version 10.1.2, using their respective built-in functions.

## 3. Results

### 3.1. Wastewater monitoring shows adenovirus outbreak

As part of the “Arizona Health Observatory” public health surveillance system, wastewater effluent was collected weekly from sixteen wastewater collection sites on a large, public university campus from January through May 2023 (Weeks 1-22) (**Fig. 1A**). 345 total samples were collected, as seven samples were not collected over the surveillance period due to difficulties at time of collection. Wastewater samples were subjected to total nucleic acid extraction (DNA and RNA) and sequenced using a hybrid-capture enrichment for a panel of 40 respiratory viruses (Illumina Respiratory Virus Enrichment Panel v2; Illumina, San Diego, CA). Sequencing reads were quality-filtered then mapped to a custom database of reference sequences (**Supplementary Fig. S1D**). Samples had a median read count of 6.6 x 10^6^ reads after quality filtering (IQR: 3.5 x 10^6^ – 9.33 x 10^6^ reads). The most abundant viruses detected were human coronaviruses (SARS-CoV-2, HCoV-NL63, HCoV-OC43, HCoV-HKU1), human adenoviruses (HAdV-B1 and HAdV-E), human influenza viruses (influenza A and B), and human rubulavirus (**Fig. 1B**).

**Figure 1:**
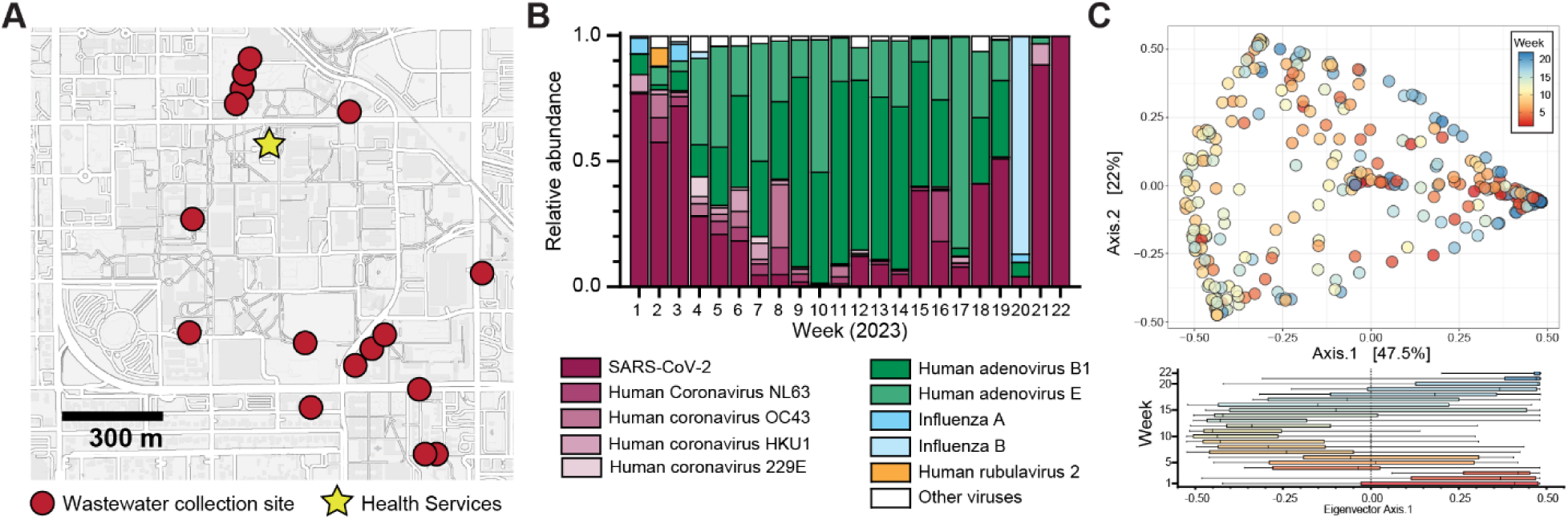
Wastewater respiratory virus surveillance on a university campus. (A) Map of university campus displaying wastewater collection sites (red circles) and the on-campus health services clinic (star). (B) Relative abundance of respiratory viruses reaching 5% abundance threshold at least one week over the 22-week surveillance period in 2023. (C) Weighted beta diversity (Bray-Curtis dissimilarity) plotted as PCoA of wastewater samples. Nodes are colored by collection week. Box plot representation of extracted eigenvector values grouped by collection week.

To investigate the viral dynamics between wastewater samples, we performed a principal coordinate analysis of weighted Bray-Curtis dissimilarity distances calculated from relative viral abundances in each sample (**Fig. 1C**). Samples collected during the mid-period (weeks 6-17) clustered distinctly suggesting that a shift in viruses occurred over this time period. This was further corroborated by Kendall’s Tau correlation tests between Axis.1 eigenvalues and SARS-CoV-2 abundance (|τ| = 0.7744) and the combined relative abundances of human adenoviruses B and E (|τ| = -0.7411 (**Fig. 1D**). Linear regression strongly modeled the inverse relationships between SARS-CoV-2 and human adenoviruses (R^2^ = 0.88 and R^2^ = 0.83, respectively) with Axis.1 Eigenvalues, whereas Axis.2 Eigenvalues were driven by an inverse relationship between HAdV-B and HAdV-E abundances (**Supplementary Figs. S1E & S1F**). Overall, the displacement of SARS-CoV-2 as the dominant wastewater respiratory virus illustrated that an outbreak of HAdV was occurring. This prompted us to investigate whether clinical presentation of HAdV increased over this time period.

### 3.2. Pathogen genome sequencing of clinical outpatients with acute respiratory infections

Given the wastewater signals, we initiated a clinical surveillance to determine if genetic signals in wastewater reflected clinical presentation. During weeks 6-21 of the wastewater surveillance period, we enrolled a cohort of 123 patients seeking care for an acute respiratory infection at the local campus Health Services clinic. The cohort was comprised of 67 female participants (mean age of 21.31 years, SD = 3.92), 52 male participants, (mean age 21.23 years, SD = 3.812), and 4 participants choosing not providing an age or gender designation. In regard to vaccination status, 69 respondents reported receiving only a SARS-CoV-2 vaccine, 2 respondents reported receiving only a current influenza vaccine, 28 respondents reported receiving both, 12 respondents reported receiving neither a SARS-CoV-2 nor current seasonal influenza vaccine, and two and nine respondents were unsure or preferred not the answer their SARS-CoV-2 or influenza vaccination status, respectively. Nasal and/or throat swab specimens were sequenced to identify the potential respiratory pathogen. Clinical sequencing samples had a median read count of 1.3 x 106 reads after quality filtering (IQR 4.0 x 10^5^ – 4.3 x 10^6^ reads).

We identified 73 viral infections from 67 participants consisting of SARS-CoV-2 (*n* = 23, 31.5%), HAdV-E (*n* = 15, 20.5%), HAdV-B (*n* = 9, 12.3%), the seasonal coronaviruses NL63 (*n* = 6, 8.2%) and HKU1 (*n* = 5, 6.8%), the human parainfluenza viruses (*n* = 8, 11.0%), metapneumovirus (*n* = 3, 4.1%), influenza viruses (*n* = 3, 4.1%), and rhinovirus (*n* = 1, 1.4%) (**Fig. 2A**). We were able to obtain high genome coverage (>90%) for 73 viral genomes (**Fig. 2B**). We identified seven cases of co-infection, with SARS-CoV-2 occurring in five of them and HAdV occurring in three. SARS-CoV-2 and HAdV-B had the greatest frequency, with identification occurring in 12 and 11 weeks out of the 16-week enrollment period, respectively.

**Figure 2:**
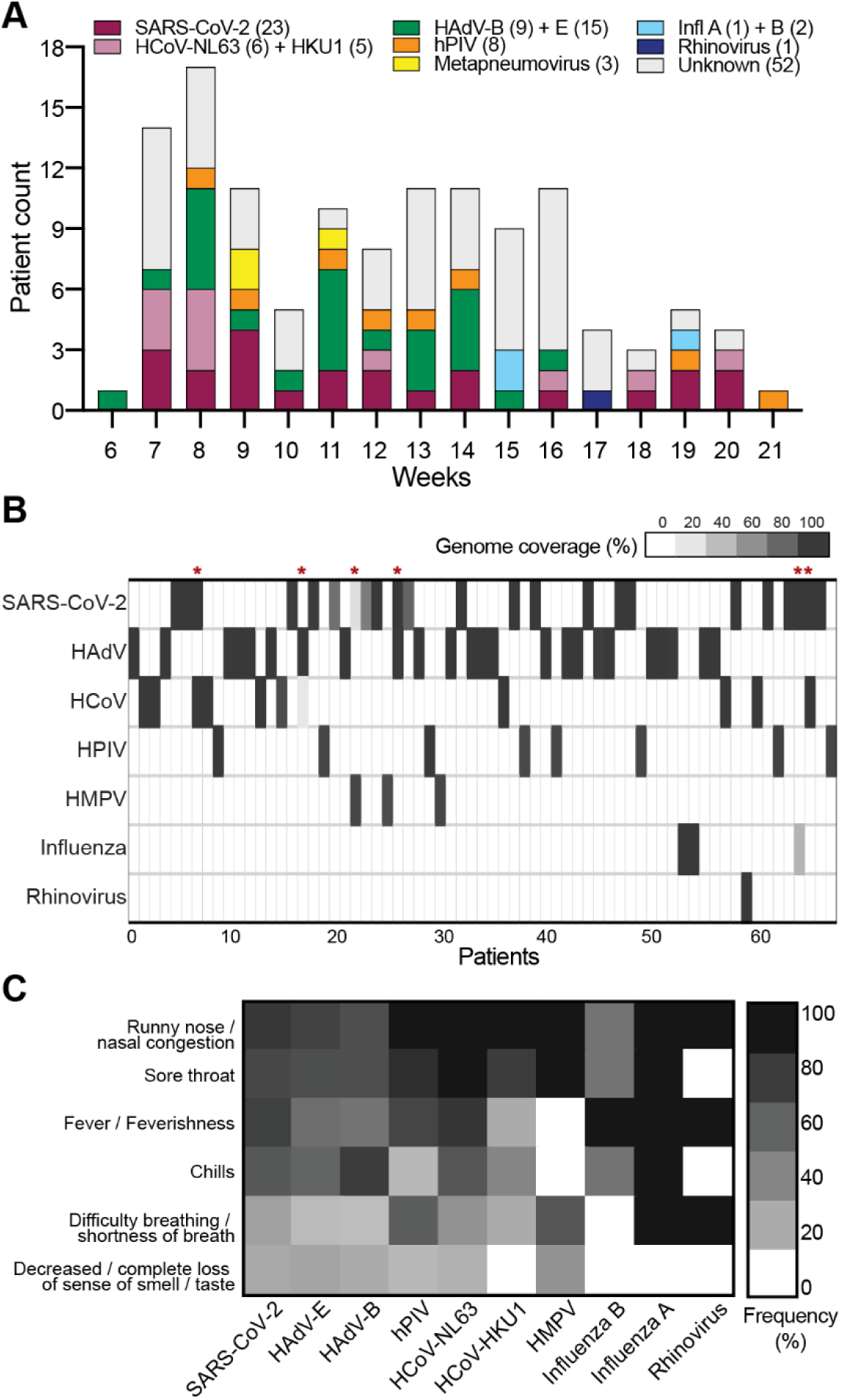
Detected viral pathogens from campus clinic patients. (A) Detected virus species over the clinic enrollment period. Numbers in parentheses indicate total number of infections over the surveillance period. (B) Heat map displaying genome coverage of viral families (y-axis) for positive patient samples (x-axis). Asterisks denote co-infections with multiple viral families. (C) Heat map displaying affirmative response rate to disease symptoms (y-axis) grouped by viral infection (x-axis).

As a primary step in diagnosis, clinical participants were polled on the presentation of symptoms. A “runny nose or nasal congestion” was the most reported symptom among respondents, followed by “sore throat,” and then “fever/feverishness.” No significant difference in responses between SARS-CoV-2 and other respiratory viruses was observed, and no evidence of a respiratory illness outbreak is indicative by symptom questionnaire responses.

### 3.3. Genomic epidemiology of human adenovirus outbreak

To determine the concordance between wastewater and clinical surveillance, we analyzed the temporal dynamics and genomic characterization of the HAdV outbreak. Viral load of HAdV was measured by qPCR using HAdV-B3, -B7, and -E4 genotype specific primers and probes (**Fig. 3A, Supplementary Figure S1G**). We observed that HAdV-E4 comprised the majority of HAdV abundance, followed by HAdV-B7. HAdv-B3 was only detected in very low levels. Lognormal distribution models estimate that HAdV-E abundance reached a maximum at week 7.53 and HAdV-B7 abundance reached a maximum at week 9.18.

**Figure 3:**
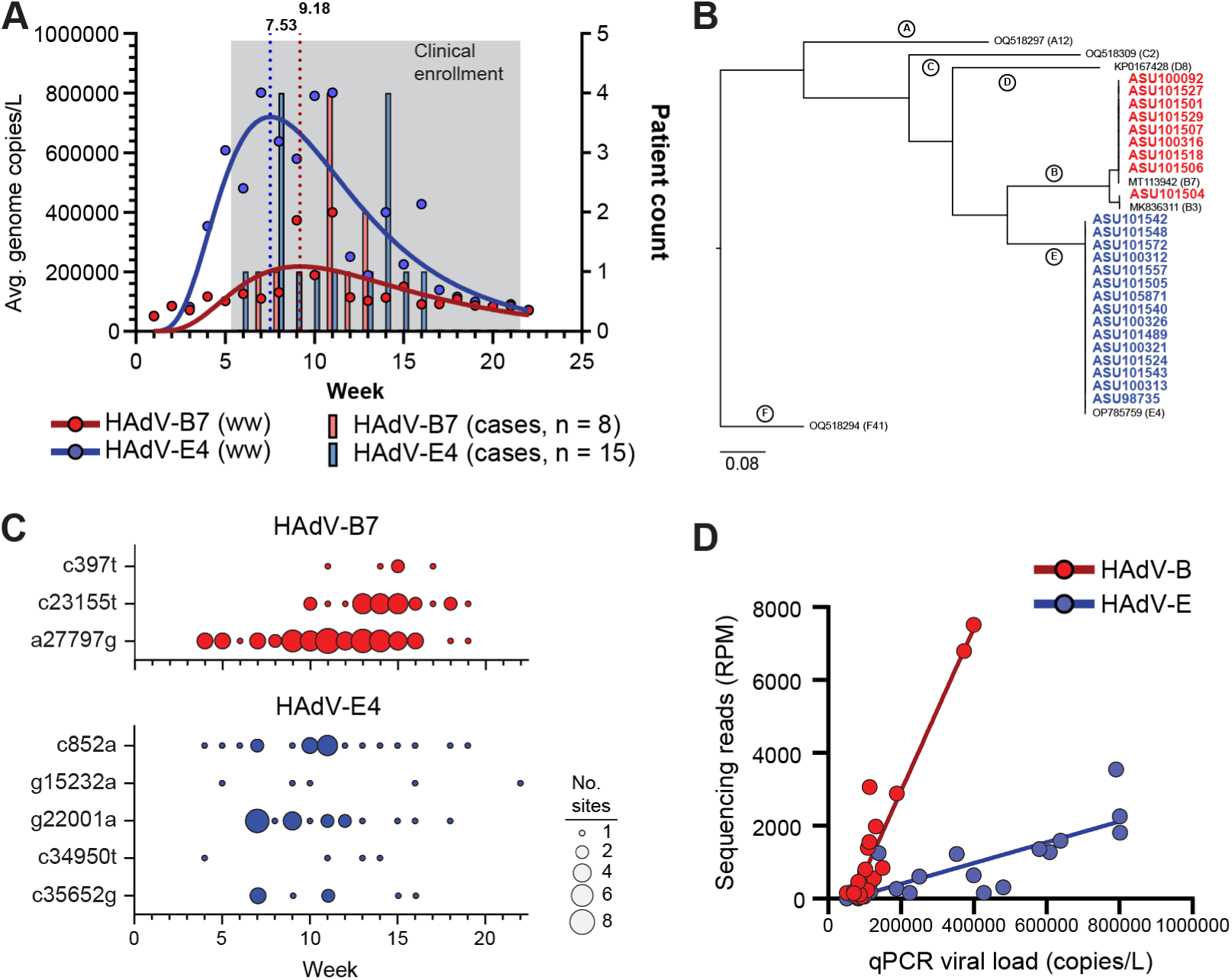
Wastewater and clinical abundance and genomic characterization of human adenoviruses B and E. (A) Left axis: Normalized viral NGS read counts (circles) and lognormal curve fits (solid lines) of wastewater samples aggregated by week. Vertical dotted lines denote the weeks of lognormal curve fit peaks. Right axis: histogram of collection week for patient samples testing positive for viral presence. (B) Whole-genome phylogenetic tree of HAdV-B (red) and HAdV-E (blue) genomes obtained from patient samples and representative Genbank samples from human adenovirus families A-F. (C) Abundance plots of wastewater samples containing the presence of HAdV minor variant SNPs. Each node displays the site (x-axis), week (y-axis), and SNP frequency of the mutation (proportion of total reads; color intensity) in the wastewater sample. (D) Average weekly qPCR-determined HAdV viral load and weekly sequencing read abundance for wastewater samples.

We found 15 clinical infections of HAdV-E4 and 8 clinical infections of HAdV-B7, corroborating the wastewater viral load composition (**Fig. 3A**). Peak clinical infections were found during weeks 8 and 14 for HAdV-E and weeks 11 and 13 for HAdV-B7. Detection of HAdV-E and HAdV-B infections stopped after weeks 16 and 13, respectively. A single HAdV-B3 infection was detected during week 11. Overall, clinical presentation of HAdV-E from this cohort was more abundant and for a longer duration than clinical presentation of HAdV-B.

Phylogenetic analysis of whole genome sequences placed all 15 HAdV-E genomes within the E4 subtype, 8 HAdV-B genomes within the B7 subtype, and 1 HAdV-B genome within the B3 subtype (**Fig. 3B**). There was no evidence of recombination, as described in certain adenoviruses (Lukashev et al., 2008). Due to the low incidence of HAdV-B3 wastewater abundance and clinical infections, we continued analysis only on the HAdV-B7 and HAdV-E4 genotypes. We found a low level of genetic divergence between our reference sequences and clinical genomes. In our clinical genomes, we detected only 3 SNPs that differed from the HAdV-B7 reference genome and 7 SNPs that differed from the HAdV-E4 reference genome (**Table 1**). There is one non-synonymous mutation in the HAdV-B7 UXP gene, c23155t (G173R substitution). There were two non-synonymous mutations in the HAdV-E4 genomes, c852a and c34950t, encoding an L95I mutation in the E1A 27 kDa protein and an E15K mutation in the E4 14.6 kDa protein. There was one mutation in a non-coding region of the B7 genomes (c397t), one mutation in a non-coding region of the E4 genomes (c35652g), and four synonymous mutations found in coding regions of the E4 genomes (g15232a, g22001a, c27053t, c27089t).

**Table 1:**
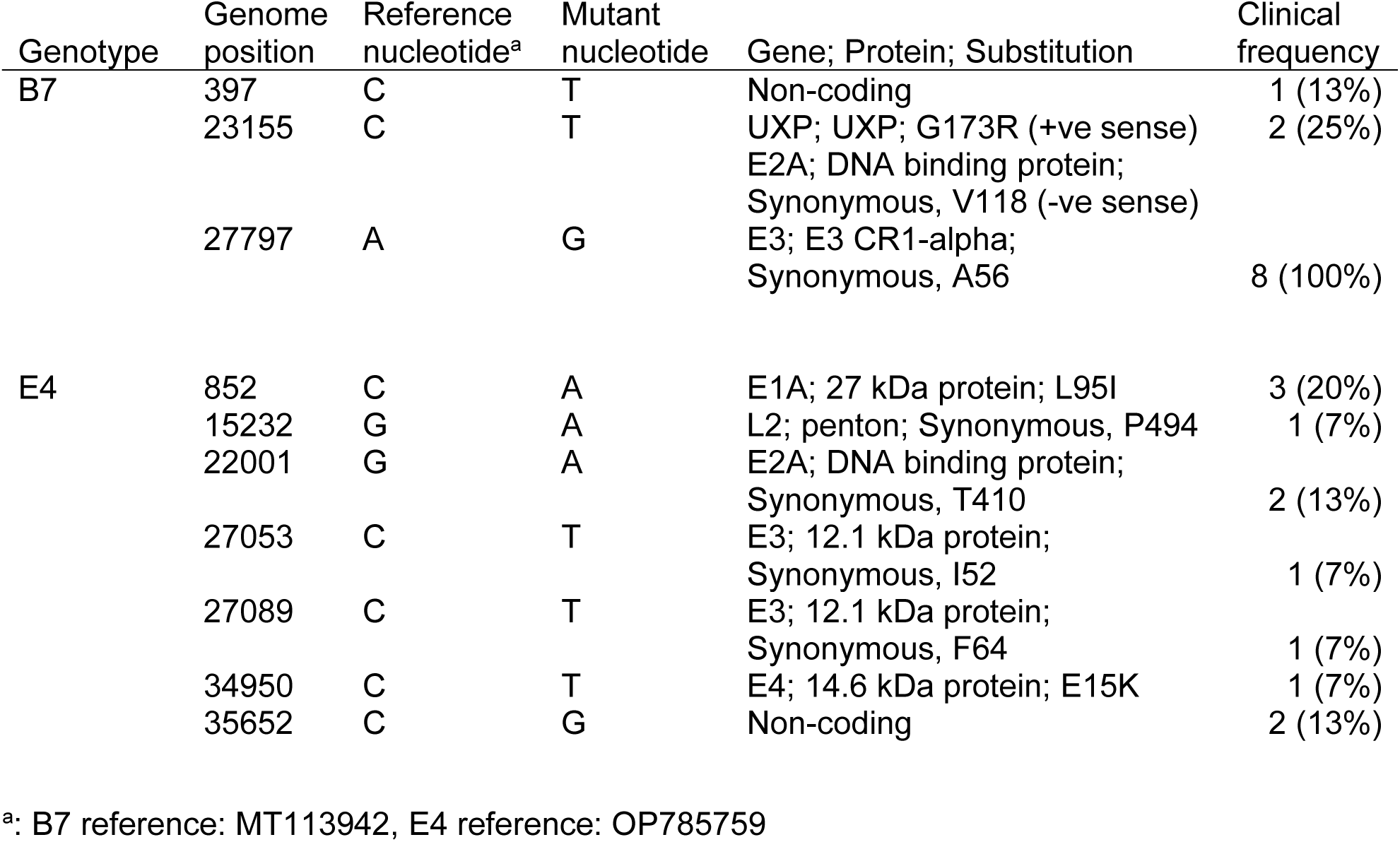
Nucleotide mutations found in HAdV-B7 and HAdV-E4 clinical specimen genomes.

We investigated whether there was genomic evidence of the clinical HAdV infections in the wastewater signals. We searched wastewater sequencing data for these mutations and detected seven of the nine SNPs found in the clinical samples (**Fig. 3C**). We were unable to detect the HAdV-E4 c27053t and HAdV-E4 c27089t mutations in the wastewater data. When we compare the patient sample collection week with the wastewater data, we observe that wastewater surveillance detected mutations on or before the week of clinical presentation, and mutations found in two or more clinical samples were found in more wastewater samples. Overall, these results indicate that the sensitivity of WBE is sufficient to track minor variant SNPs of respiratory viruses.

In order to determine if the NGS data could be used to accurately reflected viral load, we compared weekly NGS reads with viral loads measured by qPRC (**Fig 3.D**). We observed that HAdV-B sequencing reads were overrepresented compared to HAdV-E. When sequencing reads were used to estimate viral load, HAdV-B was estimated to be in greater abundance than HAdV-E (**Supplementary Figure S1H**). Lognormal models estimate viral load peak abundances at 8.21 (HAdV-E) and 9.94 (HAdV-B) weeks, which is later than qPCR estimates.

### 3.4. Genomic epidemiology of SARS-CoV-2

Wastewater SARS-CoV-2 viral load as measured by qRT-PCR was consistently detected within the surveillance period. SARS-CoV-2 (**Fig. 4A**). Hence, we compared the SARS-CoV-2 campus clinical cases to wastewater data. Phylogenetic analysis of SARS-CoV-2 genome sequences showed that most cases within this period were the recombinant XBB lineages (**Fig. 4B**). The remaining two non-XBB cases were one BQ.1 and one BA.2 descendant lineages. To investigate the relationship of SARS-CoV-2 variants circulating in the community and their presence in wastewater sequencing data, we assessed the distribution of major SARS-CoV-2 lineages in the wastewater over the study period. The fragmented nature of wastewater sequencing data makes it challenging to accurately infer sublineages from their evolutionary parents and siblings (e.g., XBB.1.5, XBB.1.5.32, and XBB.1.5.49). Hence, we aggregated sublineages to parental lineages. Consistent with the clinical variants, we observed a predominance of XBB.1.5 lineages in wastewater throughout the surveillance period. (**Fig. 4C)**. These results indicate that generally genomic SARS-CoV-2 wastewater surveillance provides information on the circulation of lineages at the community level but may lose the ability to accurately represent lineages circulating at low levels or distinguish highly similar lineages in a mixed population.

**Figure 4:**
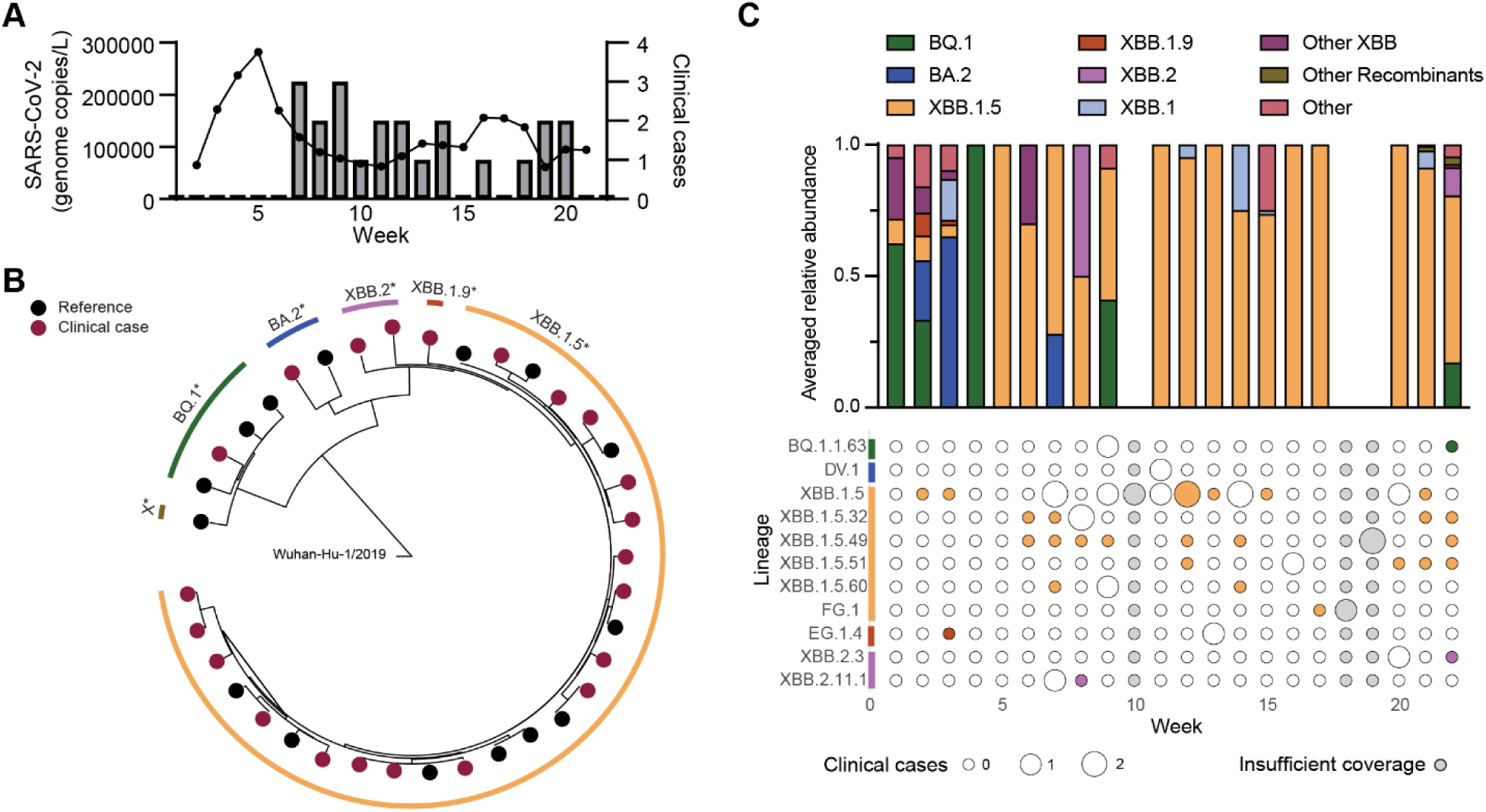
SARS-CoV-2 wastewater abundance, clinical presentation, and genomic characterization. (A) Left axis: 3 week centered average wastewater viral load in genome copies/L (line and circles) aggregated by week. Right axis: histogram of collection week for patient samples testing positive for SARS-CoV-2 presence. (B) Phylogeny of SARS-CoV-2 consensus genomes from patient samples (red circles) and references (black circles) with PANGO parental lineage colored outline. (C) Relative abundance of parental lineage-aggregated Freyja lineage estimates averaged across samples within each week (top) and bubble plot (bottom) depicting detection events of each specific lineage in the clinical cohort (bubble size) and in wastewater samples (bubble color).

### 3.5. Other prominent respiratory viruses

Finally, we sought to characterize the wastewater dynamics and phylogenetic relationships of other viruses found in the clinical cohort. Wastewater abundance models of seasonal HCoVs (HCoV-HKU1 and HCoV-NL63) and human metapneumovirus (HMPV) could be fit to lognormal distributions, whereas curve fits from the human parainfluenza virus (HPIV), influenza viruses, and human rhinovirus (HRV) were poor, likely due to low and fluctuating read abundances (**Supplementary** Fig. 2A). To understand the phylogenetic composition of viral infections, we constructed whole-genome phylogenetic trees with our clinical genomes and representative members of viral genotypes (**Supplementary** Fig. 2B). We identified five human coronavirus HKU1 genomes belonging to the A and C2 S-gene genotypes and six coronavirus NL63 genomes belonging to the B and C2 S-gene genotypes. We identified two HPIV1 genomes belonging to the 2 and 3 HN-gene genotypes, one HPIV2 genome belonging to the G3 HN-genotype, three HPIV3 genomes belonging to the C3a and C5 HN-gene genotypes, and one HPIV4 genome belonging to the 4a HN-gene genotype. We identified 3 influenza genomes belonging to the A/H1 and B/Victoria HA-gene genotypes. We identified 3 HMPV genomes belonging to the A2b2 genotype. Notably, these genomes contain the 111-base pair duplication in the G gene and may represent the first identification of this genotype in North America (Munoz-Escalante et al., 2022). Finally, we identified one HRV genome belonging to the A genotype. No evidence of large-scale recombination was observed in genomes from the clinical cohort.

## 4. Discussion

This study demonstrates the efficacy of a public health system that integrates monitoring of small-site wastewater catchments together with clinical surveillance. While treatment plant collection is economically alluring, longer *in situ* sample retention time may lead to variability in biomarker decay and changes in sampling population demographics (Amoah et al., 2020; Hart and Halden, 2020). Alternatively, distributed, small-scale sewer monitoring can improve sensitivity and narrow surveillance demographics (Yaniv et al., 2021). This study corroborates those claims, illustrating that wastewater viral surveillance reflects clinical presentation at a neighborhood level. This is one of only a few studies looking at whole-genome sequencing of respiratory viruses in both wastewater and clinical samples (Wyler et al., 2022). Development of sewer-based wastewater surveillance programs in areas of dense congregation and prone to respiratory virus outbreaks (e.g., education campuses, training grounds), could benefit these institutions by providing guidance to the treatment and prevention of these outbreaks.

Wastewater dynamics over the surveillance period are largely defined by the dynamics of SARS-CoV-2, HAdV-B, and HAdV-E. Early weeks have high SARS-CoV-2 abundance, which is displaced by HAdV, and then returns to high abundance during later weeks (**Fig. 1B**). As SARS-CoV-2 becomes increasingly endemic, displacement of SARS-CoV-2 as the majority respiratory virus in wastewater could serve as an indicator of respiratory virus outbreaks. Wastewater surveillance systems would then serve dual roles: (i) monitoring SARS-CoV-2 genomes for variants of interest (see **Fig. 4C**), and (ii) monitoring for the increased abundance of other respiratory viruses and their variants.

Within our clinical genomes only two and seven bases showed variation within B7 and E4 genotypes, respectively (**Table 1**). No amino acid variants within genotypes were detected in clinical genomes for the fiber, penton, and hexon proteins, which are commonly used as vaccine candidates (Hu et al., 2018; Vujadinovic and Vellinga, 2018). We were able to detect minor variant HAdV SNPs in wastewater signals deriving from only one or two clinical genomes (**Table 1**). Despite their low occurrence in clinical samples, the SNPs were detected at numerous wastewater sites, for extended periods of time (**Fig. 3C**). While we were unable to detect 2 clinical SNPs in the wastewater data, both of these mutations were derived from the same clinical sample.

Although wastewater qPCR measurements showed that HAdV-E viral load was greater than HAdV-B (**Fig. 3A**), HAdV-B NGS sequencing reads were approximately 60% more abundant than HAdV-E (**Fig. S1H**). Notably, there were more clinical cases of HAdV-E, and reported symptoms of HAdV-E and HAdV-B infections in this outbreak were similar (e.g., not due to milder HAdV-B infections leading to lesser outpatient visits). Taken together, and considering that the NGS approach is based on hybrid-capture enrichment, this suggests that differences in the probe-specificity between HAdV subtypes for the enrichment process may be skewing the sequencing read counts quantitatively. Whereas the qPCR assays were performed directly on nucleic acid extracts and the abundances would not be similarly skewed. Studies have shown that viral enrichment is essential to wastewater genomic sequencing given the complexity of the wastewater sample matrix (Child et al., 2023). While our analyses identified potential issues of hybrid-capture NGS, we also show how qPCR approaches can be used to supplement data interpretation as needed.

## 5. Conclusions

Here, we demonstrate the implementation of wastewater and clinical surveillance for respiratory viruses on a large, public university. We show that the wastewater viral dynamics of early 2023 was characterized by SARS-CoV-2 abundance becoming displaced by HAdV before returning to SARS-CoV-2 as the dominant virus. The increase in wastewater abundance was corroborated by the detection of a large number of HAdV infections from an on-campus clinic surveillance. Phylogenetic analysis of clinical HAdV samples revealed most genomes belonged to the E4 and B7 subtypes, with low genetic variability within subtypes. Over this time period HCoVs (SARS-CoV-2, -HKU1, -NL63), HMPV, HPIV, influenza, and HRV infections were also detected by clinical and wastewater surveillance.

## Acknowledgements

We thank Regan A. Sullins, Alexis Thomas, Michelle Tan and Gabrielle M. Hernandez Barrera for technical assistance.

## Author contributions

Conceptualization: E.S.L.; Formal analysis: S.C.H., M.F.S.; Investigation: S.C.H., M.F.S., E.M.D.; Resources: E.M.D., R.U.H., V.M.; Data curation: S.C.H., M.F.S., R.M.; Writing-original draft: S.C.H., M.F.S., E.S.L.; Writing-review and editing: S.C.H., E.S.L.; Visualization: R.M.; Supervision: E.S.L.; Funding acquisition: V.M., R.U.H., E.S.L. All authors reviewed and ap- proved the final manuscript.

## Data availability

Sequencing data has been deposited to NCBI SRA (BioProject ID PRJNA1090887). Consensus genome sequences for human adenovirus B3, human adenovirus E4, human adenovirus B7, rhinovirus A, human metapneumovirus, human parainfluenza virus 1, human parainfluenza virus 2, human parainfluenza virus 3, human parainfluenza virus 4, human coronavirus NL63, human coronavirus HKU1 have been submitted to NCBI GenBank (accession numbers pending). SARS-CoV-2 genome consensus sequences have been deposited in the GISAID database with Accession IDs: EPI_ISL_17162425, EPI_ISL_17162426, EPI_ISL_17162427, EPI_ISL_17162428, EPI_ISL_17162429, EPI_ISL_17194616, EPI_ISL_17253217, EPI_ISL_17253218, EPI_ISL_17253219, EPI_ISL_17253220, EPI_ISL_17323764, EPI_ISL_17416692, EPI_ISL_17416693, EPI_ISL_17552873, EPI_ISL_17552874, EPI_ISL_17590094, EPI_ISL_17658673, EPI_ISL_17658674, EPI_ISL_17738172, EPI_ISL_17738173, EPI_ISL_17783646, EPI_ISL_17789350. Influenza genome consensus sequences have been deposited in the GISAID database with Accession IDs: EPI_ISL_17648695 and EPI_ISL_17648783.

---

**Supplementary Figure 1:**
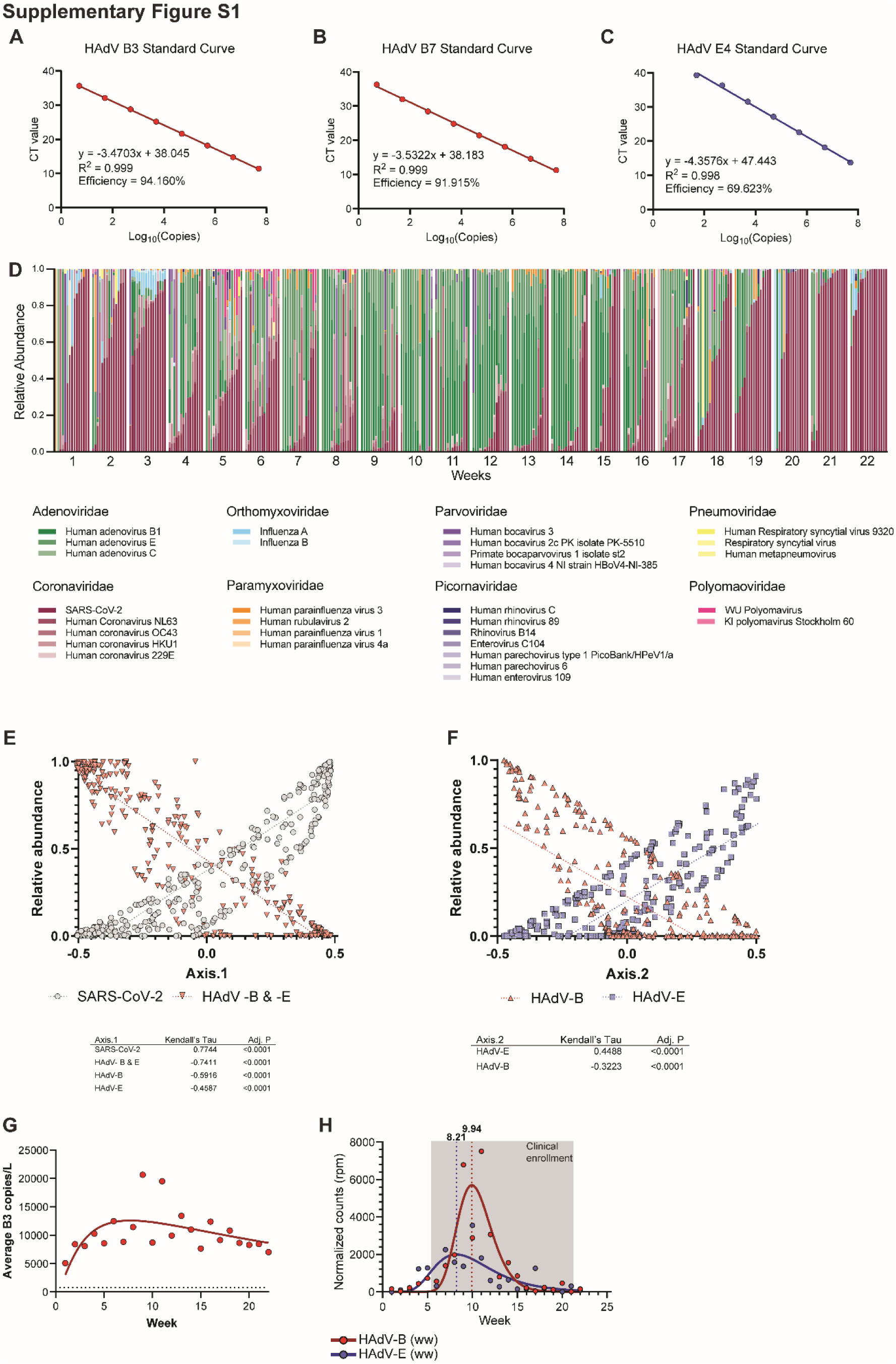
Wastewater surveillance of respiratory viruses. (A) qPCR efficiency curve of HAdV-B3 assay. (B) qPCR efficiency curve of HAdV-B7 assay. (C) qPCR efficiency curve of HAdV-E4 assay. (D) Relative abundance of viruses detected for each wastewater sample is shown. Wastewater samples were grouped by MMWR week. (E) Viral relative abundances and Axis.1 eigenvalues of wastewater samples. Table: Viruses strongly correlated (|τ|≥ 0.3) with Axis.1 eigenvalues. P values were corrected for multiple test false discovery using the Benjamini-Hochberg method. (F) Viral relative abundances and Axis.1 eigenvalues of wastewater samples. Table: Viruses strongly correlated (|τ|≥ 0.3) with Axis.2 eigenvalues. P values were corrected for multiple test false discovery using the Benjamini-Hochberg method. (G) Average weekly viral load of HAdV-B3 in wastewater samples measured by qPCR. (H) Wastewater NGS reads per week. Vertical dotted lines mark peaks of lognormal curve fits. Shaded region highlights the sampling period when clinical specimens were collected.

**Supplementary Figure 2:**
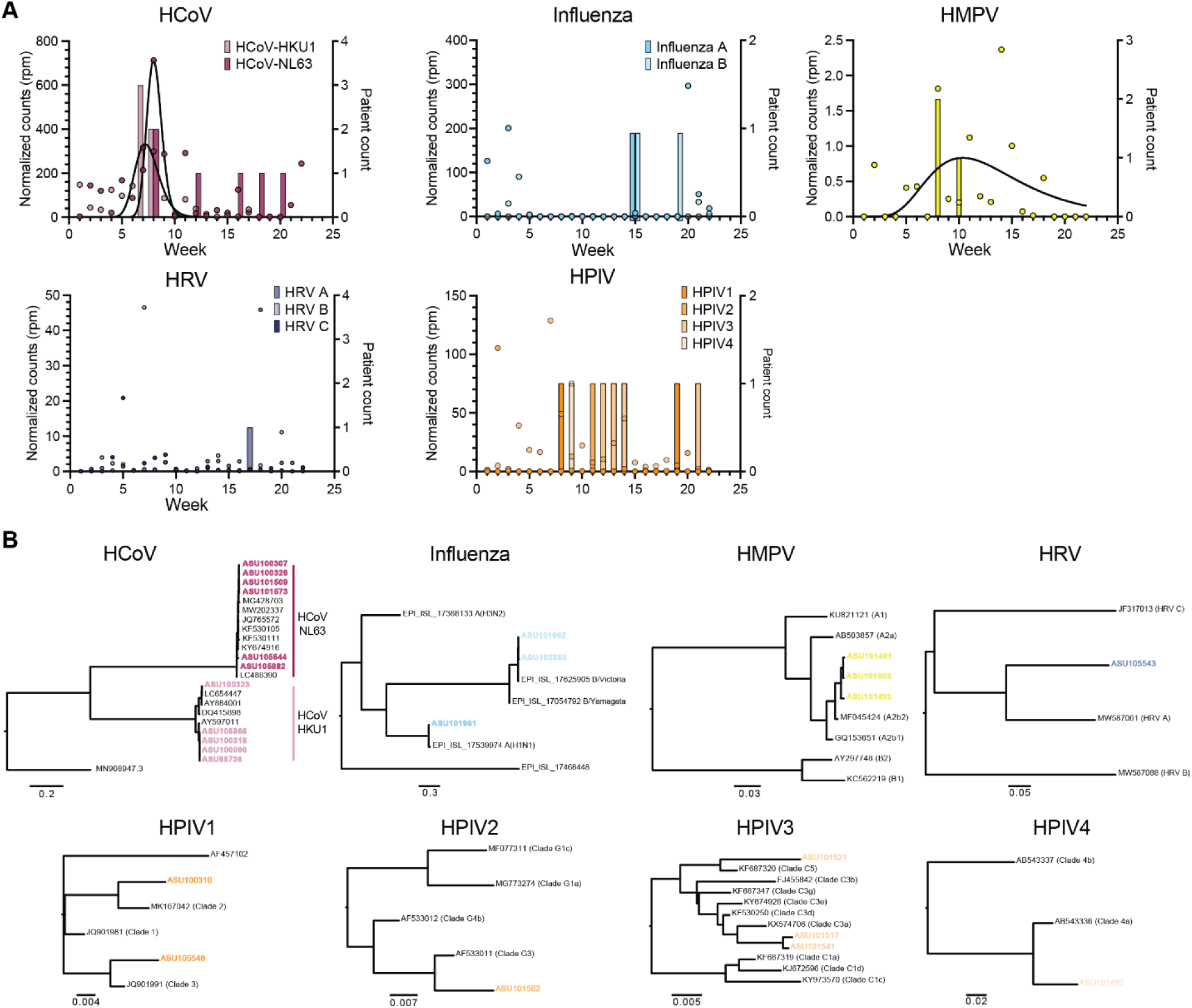
Genomic characterization of respiratory viruses. (A) Left axes: Normalized viral NGS read counts (circles) and lognormal curve fits (solid lines) of wastewater samples aggregated by week. (B) Whole-genome phylogenetic trees of virus genomes obtained from clinical patient samples (color, boldface) and representative reference sequences from NCBI GenBank or GISAID from major viral genotypes.

## References

1. Amoah ID, Kumari S, Bux F. Coronaviruses in wastewater processes: Source, fate and potential risks. Environ Int 2020; 143: 105962.

2. Arnold A, MacMahon E. Adenovirus infections. Medicine 2017; 45: 777–780.

3. Boehm AB, Hughes B, Duong D, Chan-Herur V, Buchman A, Wolfe MK, et al. Wastewater concentrations of human influenza, metapneumovirus, parainfluenza, respiratory syncytial virus, rhinovirus, and seasonal coronavirus nucleic-acids during the COVID-19 pandemic: a surveillance study. Lancet Microbe 2023; 4: e340–e348.

4. Boon H, Meinders AJ, van Hannen EJ, Tersmette M, Schaftenaar E. Comparative analysis of mortality in patients admitted with an infection with influenza A/B virus, respiratory syncytial virus, rhinovirus, metapneumovirus or SARS-CoV-2. Influenza Other Respir Viruses 2024; 18: e13237.

5. Bowes DA, Driver EM, Kraberger S, Fontenele RS, Holland LA, Wright J, et al. Leveraging an established neighbourhood-level, open access wastewater monitoring network to address public health priorities: a population-based study. Lancet Microbe 2023; 4: e29–e37.

6. Bushnell B. BBMap. 2023, 2014.

7. Castellano S, Cestari F, Faglioni G, Tenedini E, Marino M, Artuso L, et al. iVar, an Interpretation-Oriented Tool to Manage the Update and Revision of Variant Annotation and Classification. Genes (Basel) 2021; 12.

8. Cheng J, Qi X, Chen D, Xu X, Wang G, Dai Y, et al. Epidemiology and transmission characteristics of human adenovirus type 7 caused acute respiratory disease outbreak in military trainees in East China. Am J Transl Res 2016; 8: 2331–42.

9. Child HT, Airey G, Maloney DM, Parker A, Wild J, McGinley S, et al. Comparison of metagenomic and targeted methods for sequencing human pathogenic viruses from wastewater. mBio 2023; 14: e0146823.

10. Choi PM, Tscharke BJ, Donner E, O’Brien JW, Grant SC, Kaserzon SL, et al. Wastewater- based epidemiology biomarkers: Past, present and future. TrAC Trends in Analytical Chemistry 2018; 105: 453–469.

11. Fendrick AM, Monto AS, Nightengale B, Sarnes M. The economic burden of non-influenza- related viral respiratory tract infection in the United States. Arch Intern Med 2003; 163: 487–94.

12. Fontenele RS, Yang Y, Driver EM, Magge A, Kraberger S, Custer JM, et al. Wastewater surveillance uncovers regional diversity and dynamics of SARS-CoV-2 variants across nine states in the USA. Sci Total Environ 2023; 877: 162862.

13. Goka EA, Vallely PJ, Mutton KJ, Klapper PE. Single, dual and multiple respiratory virus infections and risk of hospitalization and mortality. Epidemiol Infect 2015; 143: 37–47.

14. Hadfield J, Megill C, Bell SM, Huddleston J, Potter B, Callender C, et al. Nextstrain: real-time tracking of pathogen evolution. Bioinformatics 2018; 34: 4121–4123.

15. Haramoto E, Kitajima M, Kishida N, Konno Y, Katayama H, Asami M, et al. Occurrence of pepper mild mottle virus in drinking water sources in Japan. Appl Environ Microbiol 2013; 79: 7413–8.

16. Hart OE, Halden RU. Simulated 2017 nationwide sampling at 13,940 major U.S. sewage treatment plants to assess seasonal population bias in wastewater-based epidemiology. Sci Total Environ 2020; 727: 138406.

17. Hewitt J, Greening GE, Leonard M, Lewis GD. Evaluation of human adenovirus and human polyomavirus as indicators of human sewage contamination in the aquatic environment. Water Res 2013; 47: 6750–61.

18. Hu K, Fu M, Guan X, Zhang D, Deng X, Xiao Y, et al. Penton base induces better protective immune responses than fiber and hexon as a subunit vaccine candidate against adenoviruses. Vaccine 2018; 36: 4287–4297.

19. Karthikeyan S, Levy JI, De Hoff P, Humphrey G, Birmingham A, Jepsen K, et al. Wastewater sequencing reveals early cryptic SARS-CoV-2 variant transmission. Nature 2022; 609: 101–108.

20. Katoh K, Misawa K, Kuma K, Miyata T. MAFFT: a novel method for rapid multiple sequence alignment based on fast Fourier transform. Nucleic Acids Res 2002; 30: 3059–66.

21. Li H, Durbin R. Fast and accurate short read alignment with Burrows-Wheeler transform. Bioinformatics 2009; 25: 1754–60.

22. Li H, Handsaker B, Wysoker A, Fennell T, Ruan J, Homer N, et al. The Sequence Alignment/Map format and SAMtools. Bioinformatics 2009; 25: 2078–9.

23. Linster M, Do LAH, Minh NNQ, Chen Y, Zhe Z, Tuan TA, et al. Clinical and Molecular Epidemiology of Human Parainfluenza Viruses 1-4 in Children from Viet Nam. Sci Rep 2018; 8: 6833.

24. Lion T, Baumgartinger R, Watzinger F, Matthes-Martin S, Suda M, Preuner S, et al. Molecular monitoring of adenovirus in peripheral blood after allogeneic bone marrow transplantation permits early diagnosis of disseminated disease. Blood 2003; 102: 1114–20.

25. Lowry SA, Wolfe MK, Boehm AB. Respiratory virus concentrations in human excretions that contribute to wastewater: a systematic review and meta-analysis. J Water Health 2023; 21: 831–848.

26. Lu X, Trujillo-Lopez E, Lott L, Erdman DD. Quantitative real-time PCR assay panel for detection and type-specific identification of epidemic respiratory human adenoviruses. J Clin Microbiol 2013; 51: 1089–93.

27. Lukashev AN, Ivanova OE, Eremeeva TP, Iggo RD. Evidence of frequent recombination among human adenoviruses. J Gen Virol 2008; 89: 380–388.

28. Lun JH, Crosbie ND, White PA. Genetic diversity and quantification of human mastadenoviruses in wastewater from Sydney and Melbourne, Australia. Sci Total Environ 2019; 675: 305–312.

29. Minh BQ, Schmidt HA, Chernomor O, Schrempf D, Woodhams MD, von Haeseler A, et al. IQ- TREE 2: New Models and Efficient Methods for Phylogenetic Inference in the Genomic Era. Mol Biol Evol 2020; 37: 1530–1534.

30. Moriyama M, Hugentobler WJ, Iwasaki A. Seasonality of Respiratory Viral Infections. Annu Rev Virol 2020; 7: 83–101.

31. Munoz-Escalante JC, Mata-Moreno G, Rivera-Alfaro G, Noyola DE. Global Extension and Predominance of Human Metapneumovirus A2 Genotype with Partial G Gene Duplication. Viruses 2022; 14.

32. Nao N, Saikusa M, Sato K, Sekizuka T, Usuku S, Tanaka N, et al. Recent Molecular Evolution of Human Metapneumovirus (HMPV): Subdivision of HMPV A2b Strains. Microorganisms 2020; 8.

33. Prado T, Fumian TM, Mannarino CF, Resende PC, Motta FC, Eppinghaus ALF, et al. Wastewater-based epidemiology as a useful tool to track SARS-CoV-2 and support public health policies at municipal level in Brazil. Water Res 2021; 191: 116810.

34. Shao N, Liu B, Xiao Y, Wang X, Ren L, Dong J, et al. Genetic Characteristics of Human Parainfluenza Virus Types 1-4 From Patients With Clinical Respiratory Tract Infection in China. Front Microbiol 2021; 12: 679246.

35. Shao N, Zhang C, Dong J, Sun L, Chen X, Xie Z, et al. Molecular evolution of human coronavirus-NL63, -229E, -HKU1 and -OC43 in hospitalized children in China. Front Microbiol 2022; 13: 1023847.

36. Sims N, Kasprzyk-Hordern B. Future perspectives of wastewater-based epidemiology: Monitoring infectious disease spread and resistance to the community level. Environ Int 2020; 139: 105689.

37. Smith MF, Holland SC, Lee MB, Hu JC, Pham NC, Sullins RA, et al. Baseline Sequencing Surveillance of Public Clinical Testing, Hospitals, and Community Wastewater Reveals Rapid Emergence of SARS-CoV-2 Omicron Variant of Concern in Arizona, USA. mBio 2023; 14: e0310122.

38. Tori ME, Chontos-Komorowski J, Stacy J, Lamson DM, St George K, Lail AT, et al. Identification of Large Adenovirus Infection Outbreak at University by Multipathogen Testing, South Carolina, USA, 2022. Emerg Infect Dis 2024; 30: 358–362.

39. Vujadinovic M, Vellinga J. Progress in Adenoviral Capsid-Display Vaccines. Biomedicines 2018; 6.

40. Wei J, Zang N, Zhang J, He Y, Huang H, Liu X, et al. Genome and proteomic analysis of risk factors for fatal outcome in children with severe community-acquired pneumonia caused by human adenovirus 7. J Med Virol 2023; 95: e29182.

41. Woo PC, Lau SK, Yip CC, Huang Y, Tsoi HW, Chan KH, et al. Comparative analysis of 22 coronavirus HKU1 genomes reveals a novel genotype and evidence of natural recombination in coronavirus HKU1. J Virol 2006; 80: 7136–45.

42. Wright J, Driver EM, Bowes DA, Johnston B, Halden RU. Comparison of high-frequency in-pipe SARS-CoV-2 wastewater-based surveillance to concurrent COVID-19 random clinical testing on a public U.S. university campus. Sci Total Environ 2022; 820: 152877.

43. Wyler E, Lauber C, Manukyan A, Deter A, Quedenau C, Alves LGT, et al. Comprehensive profiling of wastewater viromes by genomic sequencing. bioRxiv 2022: 2022.12.16.520800.

44. Yaniv K, Shagan M, Lewis YE, Kramarsky-Winter E, Weil M, Indenbaum V, et al. City-level SARS-CoV-2 sewage surveillance. Chemosphere 2021; 283: 131194.

45. Yusof MA, Rashid TR, Thayan R, Othman KA, Hasan NA, Adnan N, et al. Human adenovirus type 7 outbreak in Police Training Center, Malaysia, 2011. Emerg Infect Dis 2012; 18: 852–4.

